# Depression during the COVID-19 pandemic amongst residents of homeless shelters in France

**DOI:** 10.1101/2021.04.23.21255993

**Authors:** Honor Scarlett, Camille Davisse-Paturet, Cécile Longchamps, Tarik El Aarbaoui, Cécile Allaire, Anne-Claire Colleville, Mary Convence-Arulthas, Lisa Crouzet, Simon Ducarroz, Maria Melchior, the ECHO study group

## Abstract

**Background:** Accumulating evidence suggests that the COVID-19 pandemic has negatively affected global mental health and well-being. However, the impact amongst homeless persons has not been fully evaluated. The ECHO study reports factors associated with depression amongst the homeless population living in shelters in France during the Spring of 2020.

**Methods:** Interview data were collected from 527 participants living in temporary and/or emergency accommodation following France’s first lockdown (02/05/20 – 07/06/20), in the metropolitan regions of Paris (74%), Lyon (19%) and Strasbourg (7%). Interviews were conducted in French, English, or with interpreters (33% of participants, ∼20 languages). Presence of depression was ascertained using the Patient Health Questionnaire (PHQ-9).

**Results:** Amongst ECHO study participants, 30% had symptoms of moderate to severe depression (PHQ-9≥ 10). Multivariate analysis revealed depression to be associated with being female (aOR: 2.15; CI: 1.26-3.69), being single (aOR: 1.60; CI: 1.01-2.52), having a chronic illness (aOR: 2.32; CI: 1.43:-3.78), facing food insecurity (aOR: 2.12; CI: 1.40-3.22) and participants’ region of origin. Persons born in African and Eastern Mediterranean regions showed levels of depression comparable to those of French participants (30-33%) but higher than migrants from European countries (14%). Reduced rates of depression were observed amongst participants aged 30-49 (aOR: 0.60; CI: 0.38-0.95) and over 50 (aOR: 0.28; CI: 0.13-0.64), compared to 18-29-year-olds.

**Conclusions:** Our results indicate high levels of depression among homeless persons during the COVID-19 pandemic. The value of these findings extends beyond the health crisis, as predicted future instability and economic repercussions could particularly impact the mental health of this vulnerable group.

## Introduction

The instability and poor living conditions of both homeless persons and migrants are known risk factors for depression, with rates higher than in the general population [1-4]. Although estimates vary considerably, studies have found the prevalence of depression within homeless populations to range from 11-58% [5], with figures amongst migrants dependent on the host country [6], time since arrival [4] and reason for departure [7]. Moreover, the number of homeless persons [8, 9] and the proportion of migrants among them [10, 11] is increasing in France as in other European countries, with the forecasted economic recession following the pandemic likely to only accentuate this further [12].

Unstable housing negatively impacts mental health both directly and indirectly. Influences extend from structural problems, such as crowding and poor lighting [13-15], to social isolation and a lack of social support [16], feelings of unsafety [17-19], social stigma and a lack of control [18]. The relationship between homelessness and depression is also partly bidirectional, with mental health problems precipitating social exclusion and financial insecurity [16]. When combined with the challenges of migration, such as leaving family and loved ones, as well as difficulties in cultural integration which many migrants experience, even fewer supporting factors for good mental health remain [20].

Within the general population, mental health is of particular concern in the context of the COVID-19 pandemic, with research showing increased levels of depression [21-24], anxiety [25], post-traumatic stress [26] and sleep problems [27]. Amongst homeless persons, risk factors for depression may have been exacerbated during the health crisis, however currently available data on this issue are scarce. In the present study, we examine the prevalence of depression and associated risk factors amongst persons living in homeless shelters and temporary accommodation across the metropolitan areas of Paris, Lyon and Strasbourg during the Spring of 2020, a large majority of whom were migrant.

## Methods

### Study design

The ECHO study is a cross-sectional investigation conducted from May to June (02/05/20 – 07/06/20) amongst persons living in temporary or emergency housing following the first lockdown period in France (17/03/2020 – 10/05/2020). Centres used for recruitment were located in the regions of Paris (n = 12), Lyon (n = 5) and Strasbourg (n = 1). Interviews were conducted both in person (98%) or by telephone (2%), in French, English or participants’ chosen language, with the help of independent interpreters to minimise bias due to language barriers (33% of total sample). Over 20 languages were used, most frequently Arabic, Pashto and Dari. Participants were excluded if aged under 18 years, significantly inebriated or presenting cognitive disorders that prevented consent. The study protocol was approved by the Ethical Research Committee of the University of Paris (CER-2020-41).

### Sample

Overall, 929 persons were invited to take part in the ECHO study. Amongst them, 58% (535) agreed to participate, 28% were unavailable and 14% refused. Participants with insufficient depression (PHQ-9) data (1%) were also excluded from the following analyses.

### Assessment of depression

To determine symptoms of depression, the nine-item Patient Health Questionnaire (PHQ-9), validated for use in multicultural settings [28, 29], was used. Subjects were asked to report the frequency of their symptoms over the preceding two weeks, rated via a 4-part Likert scale. Participants’ depression score was calculated via the sum of responses to all 9 items. As supported by previous literature [30], a cut-off score of 10 was used to define a depressed (PHQ-9 ≥10) group for subsequent analyses.

### Relevant variables

Potential risk factors of depression included in the analyses were the following: age (18-29; 30-49; 50 years or more), sex (male; female), partnership status (stable partner; single), family status (no children; currently living with children; has children but living separately), highest completed education level, employment (none; only before lockdown; both before and during lockdown), region of origin based on the World Health Organization categories [31] (Africa; Eastern Mediterranean; America; South-East Asian; Western Pacific; Europe excluding France; France), duration of stay in France (<6 months; 6-12 months; 1-3 years; 3-5 years; 5+ years, including French natives), French language aptitude (low; moderate; fluent), administrative status (French native; residence permit holder; asylum seeker; no residence permit; other), health insurance (yes; no), chronic illness (yes; no), food insecurity (yes; no), feelings of safety (yes; no), exposure to theft or assault (yes; no), contact with friends/family (yes; no), and participants previous accommodation (other centre/association; unestablished shelter (e.g. camps, squats); street; friends/ family/ other).

French language aptitude was calculated from the sum score of self-reported French speaking, reading and writing ability, each rated on a 4-part Likert scale. Alongside this, participants degree of loneliness was measured based on the UCLA loneliness scale [32]. In supplementary analyses, we aimed to describe associations between depression and worries surrounding: coronavirus in general; becoming ill; friends or family falling ill; social isolation; job insecurity; complications with administrative procedures; future uncertainty; and, if unwell, being rejected or being unable to receive treatment. Participants were also questioned on their willingness to cooperate with the following preventative measures: visiting a doctor; isolating if unwell; respecting another lockdown; and getting vaccinated.

### Data analysis

To study associations between participants’ characteristics and the likelihood of depression, generalised logistic regression models were used. Participants from American (n = 6), South-East Asian (n = 5), and Western Pacific (n = 1) regions were excluded due to insufficient sample size, resulting in a final sample of 515 participants. Variables relevant for inclusion in the multivariate statistical model were determined via univariate Chi-square (X^2^) analysis, as supported by Hosmer and Lemeshow [33, 34]. A 75% confidence limit was used, based on Bursac et al. [35], to prevent the arbitrary exclusion of important variables [36, 37]. Additionally, centre type was included as a random effect in all models. Highly collinear variables were removed based on associations at 99% significance, and subsequently checked using Variance Inflation Factors (VIF) with a limit of VIF < 3.

Missing covariate values were imputed using Multivariate Imputation by Chained Equations (MICE) [38]. Post-hoc X^2^ analysis was performed with 95% confidence intervals. All data analysis was performed on R Version 4.0.3.

## Results

### Demographics

Participants’ demographic characteristics are shown in **Table 1**. Participants interviewed were primarily non-French (89%), male (76%), had been in France for less than 3 years (72%), were unemployed (71%), single or without a stable partner (61%), and scored low (vs moderate or high) on ratings for French language aptitude (54%). Roughly half (54%) of study participants had children, although only 19% were currently living with at least one them.

**Table 1.** Characteristics of ECHO study participants according to depression status (PHQ-9 score ≥ 10). France, n = 527, May-June 2020. Chi-square and p-value.

|  | Non-depressed<br>(%, n = 371) | Depressed<br>(%, n = 156) | p |
| --- | --- | --- | --- |
| Sex |  |  |  |
| Male | 72.3 (289) | 27.8 (111) | 0.10 |
| Female | 64.6 (82) | 35.4 (45) |  |
| Age |  |  |  |
| 18-29 | 65.1 (151) | 34.9 (81) | * |
| 30-49 | 72.3 (159) | 27.7 (61) |  |
| 50+ | 81.3 (61) | 18.7 (14) |  |
| Partnership status |  |  |  |
| No stable partner | 67.1 (216) | 32.9 (106) | * |
| Yes, has a stable partner | 77.6 (142) | 22.4 (41) |  |
| Family status |  |  |  |
| No children | 71.0 (174) | 29.0 (71) | 0.75 |
| Currently living with children | 75.0 (75) | 25.0 (25) |  |
| Not living with children | 71.9 (105) | 28.1 (41) |  |
| Highest education level |  |  |  |
| No school/ incomplete primary education | 72.1 (106) | 27.9 (41) | 0.48 |
| Primary or high school education | 72.0 (144) | 28.0 (56) |  |
| College/ Higher education | 66.9 (113) | 33.1 (56) |  |
| Employment status |  |  |  |
| Unemployed | 69.4 (258) | 30.6 (114) | 0.20 |
| Employed before lockdown | 72.5 (74) | 27.5 (28) |  |
| Employed before and during lockdown | 83.3 (30) | 16.7 (6) |  |
| Perceived food insecurity |  |  |  |
| Food insecure | 60.9 (123) | 39.1 (79) | *** |
| Not food insecure | 76.8 (242) | 23.2 (73) |  |
| Accommodation before lockdown |  |  |  |
| Other centre/charity | 68.6 (70) | 31.4 (32) | 0.90 |
| Unestablished shelter/squat | 68.8 (88) | 31.3 (40) |  |
| Street | 71.6 (154) | 28.4 (61) |  |
| Friends/family/other | 72.0 (59) | 28.0 (23) |  |
| Region of birth |  |  |  |
| French native | 68.4 (39) | 31.6 (18) | 0.07 |
| Africa | 67.0 (135) | 33.0 (66) |  |
| Eastern Mediterranean<sup>1</sup> | 70.4 (141) | 29.6 (59) |  |
| Europe (other than France) | 86.0 (49) | 14.0 (8) |  |
| Other | 58.3 (7) | 41.7 (5) |  |
| Administrative status |  |  |  |
| French native | 68.4 (39) | 31.6 (18) | 0.25 |
| Residence permit | 65.9 (93) | 34.1 (27) |  |
| Asylum seeker | 77.5 (110) | 22.5 (57) |  |
| No residence permit | 75.0 (92) | 25.0 (43) |  |
| Other | 68.1 (33) | 31.9 (11) |  |
| **Health status** |  |  |  |
| Chronic illness (yes) | 61.2 (82) | 38.8 (52) | ** |
| Chronic illness (no) | 73.8 (281) | 26.2 (100) |  |
| **Healthcare** |  |  |  |
| Medically insured/covered<sup>2</sup> | 73.1 (258) | 26.9 (95) | * |
| Uninsured | 64.7 (112) | 35.3 (61) |  |
| **French aptitude (self-reported)** |  |  |  |
| Fluent | 66.1 (76) | 33.9 (39) | 0.47 |
| Moderate | 73.0 (89) | 27.0 (33) |  |
| Low | 71.2 (203) | 28.8 (82) |  |
| **Duration of stay in France** |  |  |  |
| < 6 months | 67.8 (101) | 32.2 (48) | 0.78 |
| 6 months - 1 year | 73.6 (53) | 26.4 (19) |  |
| 1 - 3 years | 69.8 (74) | 30.2 (32) |  |
| 3 - 5 years | 66.0 (35) | 34.0 (18) |  |
| 5+ years or French native | 73.0 (100) | 27.0 (37) |  |
| **Loneliness** |  |  |  |
| Currently lonely | 63.2 (227) | 36.8 (132) | *** |
| Not currently lonely | 85.9 (140) | 14.1 (23) |  |
| **Loneliness since lockdown** |  |  |  |
| Increase in loneliness | 59.3 (115) | 40.7 (79) | *** |
| No increase in loneliness | 76.8 (252) | 23.2 (76) |  |
| **Social contact** |  |  |  |
| In regular contact with friends and family | 70.8 (323) | 29.2 (133) | 0.79 |
| No contact with friends and family | 69.2 (45) | 30.8 (20) |  |
| **Safety** |  |  |  |
| Felt unsafe since lockdown | 62.3 (96) | 37.7 (58) | * |
| Has not felt unsafe since lockdown | 73.4 (268) | 26.6 (97) |  |
| **Exposure to crime** |  |  |  |
| Exposed to theft or assault since lockdown | 67.7 (37) | 32.3 (25) | * |
| No exposure to theft or assault | 72.3 (324) | 27.7 (124) |  |

<sup>1</sup>based on WHO categories [31] (Eastern Mediterranean countries relevant to this sample: Afghanistan, Iran, Iraq, Libya, Morocco, Pakistan, Palestine, Saudi Arabia, Somalia, Sudan, Tunisia) <sup>2</sup>Including State Medical Assistance (AME) for undocumented migrants. p-value scores: <0.05 = \*, <0.01 = \*\*, <0.001 = \*\*\*

The severity of depression in the ECHO sample is shown in **Figure 1**. Less than half of participants (42%) showed no symptoms of depression, 28% had mild symptoms, 17% had moderate symptoms, 10% had moderately severe symptoms and 3% had severe symptoms.

**Figure 1.**
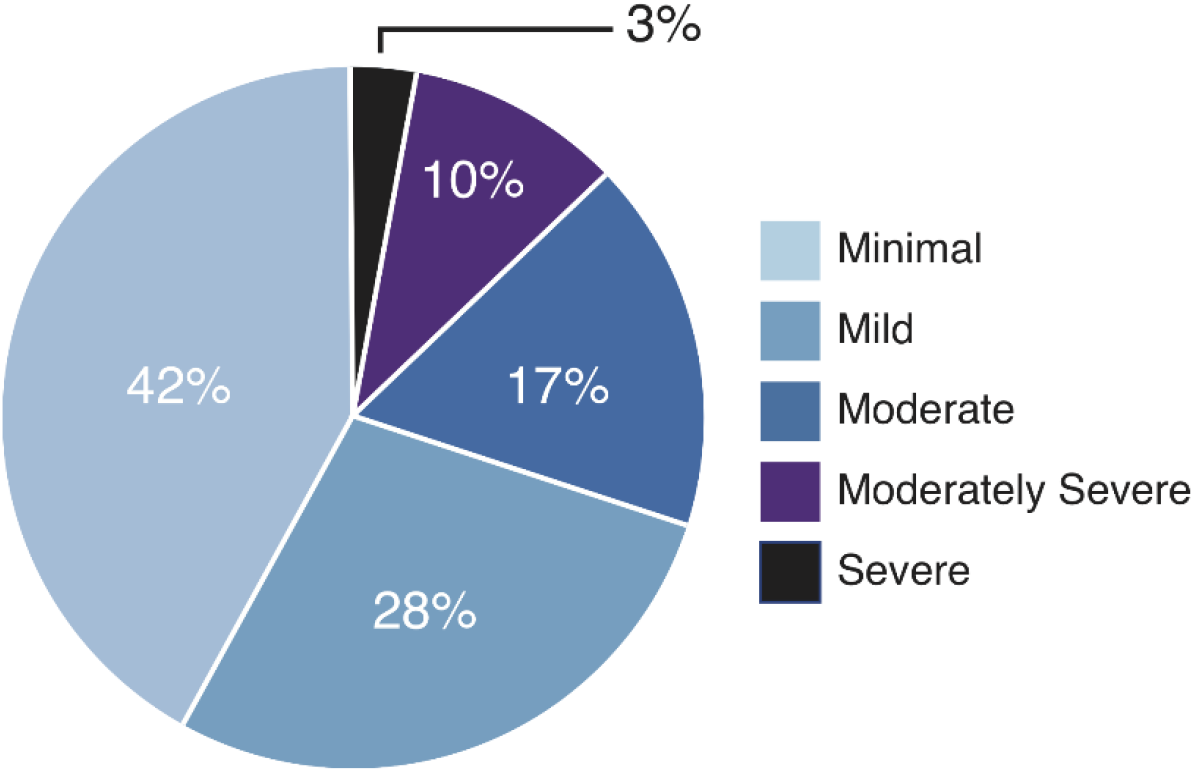
The severity of depression amongst ECHO study participants during the Spring 2020 (n = 527), based on the Patient Health Questionnaire (PHQ-9). Depression categories: Minimal = 0-4; Mild = 5-9; Moderate = 10-14; Moderately Severe = 15-19; Severe 20-27.

### Factors associated with depression

In binary analyses, characteristics associated with depression were being female (p = 0.1), young (p < 0.05), or without a stable partner (p < 0.05) and currently experiencing unemployment (p = 0.2), chronically illness (p < 0.01), food insecurity (p < 0.001) and feelings of unsafety (p < 0.05). Association was also seen for exposure to theft or assault since lockdown (p < 0.05), alongside participants region of origin (p = 0.07), administrative status (p = 0.25) and medical insurance status (p < 0.05).

Collinearity tests revealed significant associations between a) administrative status and both employment and medical insurance, as well as b) lack of safety and exposure to theft/assault. Therefore, only participants’ administrative status and lack of safety were retained for the final model.

As shown in **Table 2**, in a multivariate regression model, being female (aOR: 2.15; 95% CI: 1.26-3.69), chronically ill (aOR: 2.32; 95% CI: 1.43: 3.78), food insecure (aOR: 2.12; 95% CI: 1.40-3.22) or without a stable partner (aOR: 1.60; 95% CI: 1.01-2.52) were risk factors for depression. Lower rates of depression were seen amongst those aged 30-49 (aOR: 0.60; CI: 0.38-0.95) and 50+ (aOR: 0.28; 95% CI: 0.13-0.64), when compared to ages 18-29 years.

**Table 2.** Factors associated with depression (PHQ-9 ≥10) amongst adults living in temporary and/or emergency accommodation during the initial French lockdown period: ECHO, May-June 2020 (n = 515). Multivariate logistic regression

|  |  | n | aOR | CI |
| --- | --- | --- | --- | --- |
| **Sex** | Male | 391 | 1 |  |
|  | Female | 124 | 2.15 | 1.26-3.69 |
| **Age group** | 18-29 | 229 | 1 |  |
|  | 30-49 | 216 | 0.60 | 0.38-0.95 |
|  | 50+ | 70 | 0.28 | 0.13-0.64 |
| **Partnership status** | Has a stable partner | 185 | 1 |  |
|  | No stable partner | 330 | 1.60 | 1.01-2.52 |
| **Chronic illness** | No | 384 | 1 |  |
|  | Yes | 131 | 2.32 | 1.43-3.78 |
| **Food insecurity** | No | 311 | 1 |  |
|  | Yes | 204 | 2.12 | 1.40-3.22 |
| **Region of origin** | France | 57 | 1 |  |
|  | Africa | 201 | 0.71 | 0.31-1.62 |
|  | Eastern Mediterranean<sup>1</sup> | 200 | 0.70 | 0.30-1.63 |
|  | Europe (ex. France) | 57 | 0.25 | 0.08-0.75 |
| **Administrative status** | Residence permit<sup>2</sup> | 172 | 1 |  |
|  | Asylum seeker | 169 | 1.41 | 0.79-2.52 |
|  | No residence permit | 131 | 1.15 | 0.61-2.16 |
|  | Other | 43 | 0.98 | 0.40-2.35 |
| **Lack of safety** | No | 362 | 1 |  |
|  | Yes | 153 | 1.44 | 0.93-2.23 |

<sup>1</sup>Based on WHO categories [31] (Eastern Mediterranean countries relevant to this sample: Afghanistan, Iran, Iraq, Libya, Morocco, Pakistan, Palestine, Saudi Arabia, Somalia, Sudan, Tunisia) <sup>2</sup>Including French citizenship. aOR: Adjusted odds ratio; CI: 95% Confidence Interval.

Compared to French participants, the rate of depression was significantly lower amongst non-French Europeans (aOR: 0.25; 95% CI: 0.08-0.75). Regarding administrative status, compared to persons who were French or had a residence permit, both asylum seekers (aOR: 1.41; 95% CI: 0.79-2.52) and participants without residence permits (aOR: 1.15; 95% CI: 0.61-2.16) had higher rates of depression, however neither were significant.

The estimated variance associated with the centre type (n = 18), was 0.01 +/- 0.07, suggesting a negligible effect on depression frequency.

### Worries regarding the pandemic and depression

Associations between rates of depression and participants feelings surrounding the pandemic are shown in **Supplementary Table 1**. Participants with symptoms of depression had higher levels of worry surrounding coronavirus (p < 0.001), getting sick (p < 0.01), being rejected if sick (p < 0.05), future uncertainty (p < 0.01), isolation (p < 0.001), and access to treatment (p < 0.05) or friends and family (p < 0.01). Worries regarding administrative procedures (p < 0.01) were also seen amongst non-French participants. In comparison to the non-depressed group, depressed participants expressed greater reluctance towards respecting future lockdowns (p < 0.01).

## Discussion

Within the ECHO study, consisting of homeless persons residing in temporary and/or emergency accommodation during the Spring of 2020 (n = 527), 30% had moderate to severe depression. Associated risk factors for depression were being female, single, having a chronic illness and facing food insecurity. Moreover, persons who were French, or originated from Africa or Eastern Mediterranean regions, had higher levels of depression than persons originating from a European country other than France. Depression was associated with multiple worries and reluctance towards future lockdowns. These findings highlight the frequency of mental health difficulties and the importance of mental health care amongst persons experiencing severe socioeconomic disadvantage, which should be accounted for in strategies aiming to address the impact of the COVID-19 pandemic in vulnerable groups. To our knowledge, this is one of the first studies on the prevalence of mental health difficulties among homeless persons in the context of the COVID-19 pandemic.

### Limitations and strengths

Several limitations which may influence our findings must be acknowledged. Firstly, our study is cross-sectional, making it impossible to establish the longitudinal course of participants’ depression or the duration of symptomology. Thus, it may be that participants were already depressed prior to the pandemic. Further analyses using longitudinal samples are necessary to understand the chronology of mental health difficulties within this population, as well as the impact of homelessness on individuals’ symptoms. Secondly, our study population is unrepresentative of France’s total homeless population, consisting exclusively of those in temporary accommodation. Persons residing in alternative living situations, such as camps, squats, or the street, were not accounted for. Our findings may therefore disproportionally represent the prevalence of depression within this vulnerable population. Recruitment also consisted primarily of persons residing within two large cities (Paris and Lyon) and surrounding suburbs. Previous research has found increased loneliness during lockdown amongst adults living in urban areas compared to rural environments [39]. For this reason, the inclusion of other regions of France would have proven interesting, particularly non-urban areas.

Nevertheless, our sample is balanced and, although based on shelters expanded as a measure against the COVID-19 pandemic, includes persons who were sheltered for both short and long periods. The variety of centres used for recruitment also provided a diverse range of family living situations compared to previous research, which has often focused on solely families [10, 40] or single persons [41].

Another strong point of this study is the use of in-person interviews, which permitted the recruitment of those without access to a computer, smartphone, or internet connection. A key benefit to conducting interviews in person is the associated increase in participant response rate [42] and the possibility to include participants with low literacy who could not complete self-reported questionnaires. Given the sensitive nature of some of the study questions, this study’s low rate of missing data (1.3%) amongst variables used is commendable. Those with lower levels of education have been found to show higher rates of item non-response to health surveys, alongside males being less likely to respond to questions on depression [43]; two factors in which this population saw a majority. The use of interpreters for those unable to respond in French or English is also likely to have greatly increased the inclusivity of our dataset. Finally, the ECHO questionnaire design benefitted from collaboration with the organisations managing the homeless shelters which were included in our study.

### Prevalence of depression

The rate of depression (30%) within the ECHO sample is higher than French national averages calculated within recent years (7-10%) [44-46] alongside more global estimates [47, 48]. However, research on the rate of depression amongst homeless populations shows figures both higher [49, 50] and lower [2, 40] than in our study. These disparities may result from differences in methodology. For example, a Parisian study on homeless mental health, of which 52% were migrants, observed a depression prevalence of 57% [51]. However, recruitment took place in a free healthcare clinic, and as prior illness is a known risk factor for depression [52, 53] this elevated prevalence of depression is not surprising. In comparison, our study consisted of persons receiving accommodation after periods of exceptional adversity, considering that 65% of our sample were staying in a camp, squat, or on the street prior to lockdown. This may be reflected in the lower rate of depression. Moreover, our data collection occurred mostly in Paris and surrounding regions (74% of participants), where homelessness often results from a lack of affordable housing and is less reflective of severe poverty [10]. Unfortunately, such differences make it hard to determine what influence the pandemic had on our findings.

Despite this, the rate of depression seen was still considerably higher than the French national average during lockdown (approximately 20%) [54]. One could argue that the rate of depression in this population might demonstrate a brief, transient stage during this period of instability. However, even after treatment, the risk of relapse for depression is high, with research suggesting a 10-80% rate of recurrent episodes (dependent on depression severity) [55-59]. Therefore, identifying risk factors for depression within this population may not only alleviate suffering during times as turbulent as the pandemic or bouts of homelessness, but further improve the chance of mental stability and good health in later life.

### Migrant status

The rate of depression amongst French natives was similar to that of African and Eastern Mediterranean participants (32%, 33% and 30% respectively), and higher than those migrating from Europe (14%). This was surprising, as previous research amongst homeless populations has found depression to be more prevalent amongst migrants than local residents [51]. Moreover, despite the “healthy migrant effect”, a recurrent finding that migrants often have better health than native residents [60], recent data also suggest that migrants may actually be more vulnerable to mental health problems that non-migrants [61]. However, the causes for homelessness are also likely to vary between migrants and local residents. Our findings may then be explained by mental health difficulties increasing the likelihood of being homeless more so within native populations. Whether these rates are accentuated by the COVID-19 pandemic, or our sample consisting of solely those experiencing homelessness, is not possible to establish from our data. However, seeing as the pandemic increased the rate of financial insecurity and unemployment [62], and therefore housing instability [63], these factors are likely to confound each other.

### Demographic characteristics

Within our sample, women showed over double the risk for depression than men. It must be noted that depression was self-reported, which has previously been found to enable gender bias due to men underreporting symptoms [64]. However, the association between sex and depression (with increased prevalence amongst women) is one of the most consistent findings amongst recent research, both in France [65] and globally [66, 67].

Contrary to prior evidence [68], older participants in our study showed lower rates of depression. Even more paradoxically, previous research has found that depression increased with age during the pandemic in association with higher levels of chronic illness in older subjects [52]. Age is a known predictor of chronic illness [69], and within our cohort chronic illness was seen to increase the risk of depression significantly. One potential explanation for our findings may be that older participants felt less impacted by the pandemic. Younger subjects may be more concerned about their career, social life and future in general. Worries surrounding the future were significantly more common among depressed (75.7%) compared to non-depressed (61.2%) participants. Further research is needed to see how the differential effects of the pandemic on age groups change with time. This is of particular relevance to migrants, as European Union statistics have found migrants to be, on average, much younger than a countries native population [70]. This was echoed by our sample, in which the average age of French nationals was 12 years older than non-French subjects.

Relationship status was also associated with depression, with single participants at greater risk. Albeit potentially linked to financial security, the positive effect of having a partner may result from associated comfort or support, thereby preventing loneliness. This association between loneliness and depression is of particular relevance to the pandemic, with lockdown measures not only increasing loneliness [71], but more severely so in those with low socioeconomic status [39, 72]. Within our population, increased loneliness since the start of lockdown was seen amongst 37% of subjects. This is considerably higher than figures from COVID-19 research on the US adult population, which saw loneliness increase from 11% in 2018 to 13.8% in April 2020 [73]. Moreover, depressed participants within our study were significantly more likely to worry about remaining isolated. This may partly explain why depressed subjects were also significantly less willing to accept another lockdown.

### Socioeconomic status (SES)

Food insecurity positively associated symptoms of depression, however job loss did not. This was surprising, as literature on mental health during the pandemic found depression to associate significantly with job loss [74]. However, the low rate of employment (27%) in our cohort is likely to account for this disparity. Recent research on homeless families in Paris found food insecurity to be a major problem, affecting 77% of parents [40]. Whilst only 39% of our cohort declared food insecurity, as temporary accommodation for both winter and COVID-19 close, this figure is likely to increase. Lower SES has also been found to associate with increased depression during the pandemic [75, 76], alongside greater levels of loneliness [39, 72] and anxiety [22]. Interestingly, several studies show parallel levels of depression between homeless populations and non-homeless cohorts with low SES [2, 77]. Together, these data contribute to evidence of a strong connection between poverty and mental health.

### Health status

Chronic illness was found to significantly increase the rate of depression, which is consistent with other data [78]. This is of particular concern for those without medical insurance (33% of the total cohort), who may face greater difficulty getting treatment for both depression and chronic illness. Alongside this, psychiatric conditions are likely to confound each other, especially amongst low-income persons [49]. Seeing as the COVID-19 pandemic has also been found to trigger both post-traumatic stress [79] and anxiety [80, 81], better targeting preventative interventions for those most at-risk for all elements of chronic ill-health will likely support the reduction of depression rates.

### Future applications

This research has applications relevant not only to the COVID-19 pandemic, but to future periods of mental health disparity resulting of immediate social or health crises. Much of the temporary accommodation this study recruited from, created in response to the pandemic, was, or will soon be, subsequently disbanded [82, 83]. The effects this may have on mental health warrant investigation. Moreover, due to data collection occurring in the very first stages of the pandemic, it is possible certain social and financial effects had not yet occurred. This may explain the lack of significant association seen between fiscal worries and depression. Further investigation within this population is needed to explore the relationship between mental health and financial disruption during later stages of the pandemic. We are currently (Spring 2021) conducting a second wave of the ECHO study in a similar population augmented with persons living on the street, to gain better understanding of long-term patterns of health within this population in relation to the COVID-19 pandemic.

## Conclusion

Moderate to severe depression was seen in almost a third of homeless persons interviewed, with women, young people, those without stable partners, and chronically unwell or food insecure persons at greatest risk. Increased loneliness was also seen in 37% of subjects since the start of lockdown, alongside higher levels of worry surrounding isolation amongst depressed participants. These findings can teach us about not only health inequalities in the context of COVID-19, but also how similar circumstances may affect the mental health of future populations with comparable disadvantage. Closer attention must be paid to those most at risk, as supporting good mental health within these communities will in turn increase the likelihood of their progression to stable housing and better living conditions in general.

## Data Availability

The data that support the findings of this study are available from the Department of Social Epidemiology, Pierre Louis Institute of Epidemiology and Public Health. Restrictions apply to the availability of these data, which were used under license for this study.

## Acknowledgements

We thank all research participants who contributed to this project, as well as ISM interpreters without whom communication with study participants would have been impossible.

---

**Supplementary Table 1.** Associations between rates of depression (PHQ-9 ≥ 10) and worries expressed amongst adults living in temporary and/or emergency accommodations during the initial French lockdown period: ECHO, May-June 2020.

|  | n | Non-depressed | Depressed | p |
| --- | --- | --- | --- | --- |
| **Worries about:** |  |  |  |  |
| Coronavirus in general | 521 | 56.7%<sup>1</sup> | 73.4%<sup>1</sup> | *** |
| Getting sick | 507 | 64.0% | 78.1% | ** |
| If sick, being rejected by people | 485 | 52.0% | 62.4% | * |
| If sick, being unable to get treatment | 487 | 38.2% | 50.3% | * |
| Family or friends getting sick | 491 | 77.3% | 82.5% | 0.20 |
| No longer seeing friends and family | 494 | 62.7% | 75.0% | ** |
| Job insecurity as a result of the pandemic | 460 | 42.8% | 52.6% | 0.05 |
| Financial instability | 480 | 46.6% | 47.5% | 0.86 |
| Complications with administrative procedures<sup>2</sup> | 492 | 73.3% | 86.5% | ** |
| Future uncertainty | 475 | 61.2% | 75.7% | ** |
| Remaining isolated | 493 | 56.8% | 75.0% | *** |
| **Willingness to:** |  |  |  |  |
| Consult a doctor and/or get tested if showing any signs of coronavirus | 511 | 92.4% | 87.7% | 0.08 |
| Isolate oneself if showing any signs of coronavirus | 510 | 91.0% | 93.5% | 0.36 |
| Respect another lockdown | 505 | 94.1% | 88.0% | * |
| Get vaccinated if a coronavirus vaccine is found<sup>3</sup> | 208 | 65.1% | 71.2% | 0.40 |

<sup>1</sup>Scored moderately or above (taken from a 5-part Likert scale) <sup>2</sup>Non-French participants only <sup>3</sup>Question added 15/05/20. p-value scores based on Chi square analysis: <0.05 = \*, <0.01 = \*\*, <0.001 = \*\*\*

## References

1. Guardia D, Salleron J, Roelandt JL, Vaiva G. [Prevalence of psychiatric and substance use disorders among three generations of migrants: Results from French population cohort]. L’Encephale. 2017;43(5): 435–43.

2. Laporte A, Vandentorren S, Détrez M-A, Douay C, Le Strat Y, Le Méner E, et al. Prevalence of mental disorders and addictions among homeless people in the greater Paris area, France. International Journal of Environmental Research and Public Health. 2018;15(2): 241.

3. Hossain MM, Sultana A, Tasnim S, Fan Q, Ma P, McKyer ELJ, et al. Prevalence of mental disorders among people who are homeless: An umbrella review. International Journal of Social Psychiatry. 2020;66(6): 528–41.

4. Foo SQ, Tam WW, Ho CS, Tran BX, Nguyen LH, McIntyre RS, et al. Prevalence of Depression among Migrants: A Systematic Review and Meta-Analysis. International journal of environmental research and public health. 2018;15(9): 1986.

5. Fazel S, Khosla V, Doll H, Geddes J. The prevalence of mental disorders among the homeless in western countries: systematic review and meta-regression analysis. PLoS Med. 2008;5(12): e225.

6. Lindert J, Ehrenstein O, Priebe S, Mielck A, Brähler E. Depression and anxiety in labor migrants and refugees – A systematic review and meta-analysis. Social science & medicine (1982). 2009;69 :246–57.

7. Heeren M, Wittmann L, Ehlert U, Schnyder U, Maier T, Müller J. Psychopathology and resident status - comparing asylum seekers, refugees, illegal migrants, labor migrants, and residents. Compr Psychiatry. 2014;55(4): 818–25.

8. Institut National de la Statistique et des Etudes Economiques (INSEE), Institut national d’études démographiques (INED). Enquête auprès des personnes fréquentant les services d’hébergement et les distributions de repas chauds. ADISP-CMH; 2012.

9. Yaouancq F, Duée M. Les sans-domicile en 2012: une grande diversité de situations. France portrait social. 2014.

10. Roze M, Melchior M, Vuillermoz C, Rezzoug D, Baubet T, Vandentorren S. Post-Traumatic Stress Disorder in Homeless Migrant Mothers of the Paris Region Shelters. International journal of environmental research and public health. 2020;17(13): 4908.

11. Pierre FTFA. Third overview of housing exclusion in Europe 2018. 2018.

12. Flaming D, Orlando A, Burns P, Pickens S. Locked Out: Unemployment and Homelessness in the COVID Economy. SSRN 2021; 3765109.

13. Lima NNR, de Souza RI, Feitosa PWG, Moreira JLdS, da Silva CGL, Neto MLR. People experiencing homelessness: Their potential exposure to COVID-19. Psychiatry Research. 2020;288: 112945.

14. Campagna G. Linking crowding, housing inadequacy, and perceived housing stress. Journal of Environmental Psychology. 2016;45: 252–66.

15. Liddell C, Guiney C. Living in a cold and damp home: frameworks for understanding impacts on mental well-being. Public Health. 2015;129(3): 191–9.

16. Suglia SF, Duarte CS, Sandel MT. Housing quality, housing instability, and maternal mental health. Journal of Urban Health. 2011;88(6): 1105–16.

17. Hernández D. Affording housing at the expense of health: exploring the housing and neighborhood strategies of poor families. Journal of Family Issues. 2016;37(7): 921–46.

18. Swope CB, Hernández D. Housing as a determinant of health equity: A conceptual model. Social Science & Medicine. 2019;243: 112571.

19. Clark C, Ryan L, Kawachi I, Canner MJ, Berkman L, Wright RJ. Witnessing community violence in residential neighborhoods: a mental health hazard for urban women. Journal of Urban Health. 2008;85(1): 22–38.

20. Pannetier J, Lert F, Jauffret Roustide M, du Loû AD. Mental health of sub-saharan african migrants: The gendered role of migration paths and transnational ties. SSM Population Health. 2017;3: 549–57.

21. Shevlin M, McBride O, Murphy J, Miller JG, Hartman TK, Levita L, et al. Anxiety, depression, traumatic stress and COVID-19-related anxiety in the UK general population during the COVID-19 pandemic. British Journal of Psychiatry. 2020;6(6): e125.

22. Jia R, Ayling K, Chalder T, Massey A, Broadbent E, Coupland C, et al. Mental health in the UK during the COVID-19 pandemic: early observations. medRxiv. 2020: 20102012.

23. Fancourt D, Steptoe A, Bu F. Trajectories of depression and anxiety during enforced isolation due to COVID-19: longitudinal analyses of 59,318 adults in the UK with and without diagnosed mental illness. medRxiv. 2020: 20120923.

24. Vizard T, Davis J, White E, Beynon B. Coronavirus and depression in adults, Great Britain: June 2020. London: Office for National Statistics; 2020.

25. Sigdel A, Bista A, Bhattarai N, Pun BC, Giri G, Marqusee H, et al. Depression, Anxiety and Depression-anxiety comorbidity amid COVID-19 Pandemic: An online survey conducted during lockdown in Nepal. medRxiv. 2020: 20086926.

26. Rossi R, Socci V, Talevi D, Mensi S, Niolu C, Pacitti F, et al. COVID-19 Pandemic and Lockdown Measures Impact on Mental Health Among the General Population in Italy. Frontiers in Psychiatry. 2020; 11(790).

27. Daly M, Sutin AR, Robinson E. Longitudinal changes in mental health and the COVID-19 pandemic: evidence from the UK Household Longitudinal Study. Psychological Medicine. 2020: 1–10.

28. Huang FY, Chung H, Kroenke K, Delucchi KL, Spitzer RL. Using the Patient Health Questionnaire-9 to measure depression among racially and ethnically diverse primary care patients. Journal of general internal medicine. 2006;21(6): 547–52.

29. Arthurs E, Steele R, Hudson M, Baron M, Thombs B. Are Scores on English and French Versions of the PHQ-9 Comparable? An Assessment of Differential Item Functioning. PloS One. 2012;7: e52028.

30. Manea L, Gilbody S, McMillan D. Optimal cut-off score for diagnosing depression with the Patient Health Questionnaire (PHQ-9): a meta-analysis. Canadian Medical Association journal. 2012;184(3): e191–E6.

31. World Health Organization. List of Member States. Geneva 2004.

32. Hughes ME, Waite LJ, Hawkley LC, Cacioppo JT. A Short Scale for Measuring Loneliness in Large Surveys: Results From Two Population-Based Studies. Research on Aging. 2004;26(6): 655–72.

33. Hosmer DW, Lemeshow S. Applied survival analysis: regression modelling of time to event data: Wiley; 2002.

34. Hosmer Jr DW, Lemeshow S, Sturdivant RX. Applied logistic regression: John Wiley & Sons; 2013.

35. Bursac Z, Gauss CH, Williams DK, Hosmer DW. Purposeful selection of variables in logistic regression. Source Code for Biology and Medicine. 2008;3(1): 17.

36. Bendel RB, Afifi AA. Comparison of stopping rules in forward “stepwise” regression. Journal of the American Statistical association. 1977;72(357): 46–53.

37. Mickey RM, Greenland S. The impact of confounder selection criteria on effect estimation. American journal of epidemiology. 1989;129(1): 125–37.

38. van Buuren S, Groothuis-Oudshoorn K. mice: Multivariate Imputation by Chained Equations in R. Journal of Statistical Software. 2011; 45(3).

39. Bu F, Steptoe A, Fancourt D. Who is lonely in lockdown? Cross-cohort analyses of predictors of loneliness before and during the COVID-19 pandemic. medRxiv. 2020: 20101360.

40. Vandentorren S, Le Méner E, Oppenchaim N, Arnaud A, Jangal C, Caum C, et al. Characteristics and health of homeless families: the ENFAMS survey in the Paris region, France 2013. European Journal of Public Health. 2016;26(1): 71–6.

41. Roederer T, Mollo B, Vincent C, Nikolay B, Llosa AE, Nesbitt R, et al. Seroprevalence and risk factors of exposure to COVID-19 in homeless people in Paris, France: a cross-sectional study. The Lancet Public Health. 2021;6(4): e202–e9.

42. Bowling A. Mode of questionnaire administration can have serious effects on data quality. Journal of Public Health. 2005;27(3): 281–91.

43. Tsiampalis T, Panagiotakos DB. Missing-data analysis: socio-demographic, clinical and lifestyle determinants of low response rate on selfreported psychological and nutrition related multi-item instruments in the context of the ATTICA epidemiological study. BMC Medical Research Methodology. 2020;20(1): 148.

44. Léon C, Chan Chee C, Du Roscoät E, Andler R, Cogordan C, Guignard R, et al. La dépression en France chez les 18–75 ans: Résultats du Baromètre santé 2017. Bulletin épidémiologique hebdomadaire. 2018: 32–3.

45. Gourier-Fréry C, Guignard R, Beck F. The current state of mental health surveillance in France. Santé Publique (Vandoeuvre-Lès-Nancy, France). 2011;23: S13–29.

46. Sapinho D, Chan-Chee C, Briffault X, Guignard R, Beck F. Mesure de l’épisode dépressif majeur en population générale: apports et limites des outils. Bulletin épidémiologique hebdomadaire. 2008;35: 313–7.

47. World Health Organization. Depression and other common mental disorders: global health estimates. World Health Organization, 2017.

48. Murray CJ, Vos T, Lozano R, Naghavi M, Flaxman AD, Michaud C, et al. Disability-adjusted life years (DALYs) for 291 diseases and injuries in 21 regions, 1990–2010: a systematic analysis for the Global Burden of Disease Study 2010. The Lancet. 2012;380(9859): 2197–223.

49. Bassuk EL, Buckner JC, Perloff JN, Bassuk SS. Prevalence of mental health and substance use disorders among homeless and low-income housed mothers. American Journal of Psychiatry. 1998;155(11):1561–4.

50. Tinland A, Boyer L, Loubière S, Greacen T, Girard V, Boucekine M, et al. Victimization and posttraumatic stress disorder in homeless women with mental illness are associated with depression, suicide, and quality of life. Neuropsychiatry Disease and Treatment. 2018;14: 2269–79.

51. Rondet C, Cornet P, Kaoutar B, Lebas J, Chauvin P. Depression prevalence and primary care among vulnerable patients at a free outpatient clinic in Paris, France, in 2010: results of a cross-sectional survey. BMC Family Practice. 2013;14(1): 151.

52. Chandola T, Kumari M, Booker CL, Benzeval MJ. The mental health impact of COVID-19 and pandemic related stressors among adults in the UK. medRxiv. 2020: 20146738.

53. Goodwin GM. Depression and associated physical diseases and symptoms. Dialogues in clinical neuroscience. 2006;8(2):259–65.

54. Santé Publique France. CoviPrev: A survey to track changes in behavior and mental health during the COVID-19 epidemic. medRxiv 2021.

55. Kumagai N, Tajika A, Hasegawa A, Kawanishi N, Horikoshi M, Shimodera S, et al. Predicting recurrence of depression using lifelog data: an explanatory feasibility study with a panel VAR approach. BMC Psychiatry. 2019;19(1): 391.

56. Köhler S, Wiethoff K, Ricken R, Stamm T, Baghai TC, Fisher R, et al. Characteristics and differences in treatment outcome of inpatients with chronic vs. episodic major depressive disorders. J Affect Disord. 2015;173:126–33.

57. Holma KM, Holma IA, Melartin TK, Rytsälä HJ, Isometsä ET. Long-term outcome of major depressive disorder in psychiatric patients is variable. The Journal of clinical psychiatry. 2008;69(2):196–205.

58. Kessler RC, Bromet EJ. The epidemiology of depression across cultures. Annual review of public health. 2013;34: 119–38.

59. Limosin F, Loze J-Y, Zylberman-Bouhassira M, Schmidt ME, Perrin E, Rouillon F. The Course of Depressive Illness in General Practice. The Canadian Journal of Psychiatry. 2004;49(2): 119–23.

60. Puschmann P, Donrovich R, Matthijs K. Salmon Bias or Red Herring? : Comparing Adult Mortality Risks (Ages 30-90) between Natives and Internal Migrants: Stayers, Returnees and Movers in Rotterdam, the Netherlands, 1850-1940. Human nature (Hawthorne, NY). 2017;28(4): 481–99.

61. Aldridge RW, Nellums LB, Bartlett S, Barr AL, Patel P, Burns R, et al. Global patterns of mortality in international migrants: a systematic review and meta-analysis. The Lancet. 2018;392(10164): 2553–66.

62. Johnson MT, Johnson EA, Webber L, Nettle D. Mitigating social and economic sources of trauma: The need for universal basic income during the coronavirus pandemic. US: Educational Publishing Foundation; 2020. p. S191–S2.

63. Albon D, Soper M, Haro A. Potential implications of the COVID-19 pandemic on the homeless population. Chest. 2020;158(2): 477–8.

64. Sigmon ST, Pells JJ, Boulard NE, Whitcomb-Smith S, Edenfield TM, Hermann BA, et al. Gender Differences in Self-Reports of Depression: The Response Bias Hypothesis Revisited. Sex Roles. 2005;53(5): 401–11.

65. Fond G, Lancon C, Auquier P, Boyer L. Prévalence de la dépression majeure en France en population générale et en populations spécifiques de 2000 à 2018 : une revue systématique de la littérature. La Presse Médicale. 2019;48(4): 365–75.

66. Muñoz M, Crespo M, Pérez-Santos E. Homelessness Effects on Men’s and Women’s Health. International Journal of Mental Health. 2005;34(2): 47–61.

67. World Health Organization. The global burden of disease: 2004 update: World Health Organization; 2008.

68. Arias de la Torre J, Vilagut G, Ronaldson A, Dregan A, Ricci-Cabello I, Hatch SL, et al. Prevalence and age patterns of depression in the United Kingdom. A population-based study. Journal of Affective Disorders. 2021;279: 164–72.

69. Divo MJ, Martinez CH, Mannino DM. Ageing and the epidemiology of multimorbidity. European Respiratory Journal. 2014;44(4): 1055.

70. Eurostat - European Commission. Migration and migrant population statistics. Luxembourg: Eurostat [Internet]. 2020.

71. Bu F, Steptoe A, Fancourt D. Loneliness during lockdown: trajectories and predictors during the COVID-19 pandemic in 35,712 adults in the UK. medRxiv. 2020: 20116657.

72. Burchell B, Wang S, Kamerāde D, Bessa I, Rubery J. Cut hours, not people: no work, furlough, short hours and mental health during COVID-19 pandemic in the UK. 2020.

73. McGinty EE, Presskreischer R, Han H, Barry CL. Psychological distress and loneliness reported by US Adults in 2018 and April 2020. JAMA. 2020;324(1):93–4.

74. Posel D, Oyenubi A, Kollamparambil U. Job loss and mental health during the COVID-19 lockdown: Evidence from South Africa. PLOS ONE. 2021; 16(3): e0249352.

75. Xue B, McMunn A. Gender differences in the impact of the Covid-19 lockdown on unpaid care work and psychological distress in the UK. 2021; 16(3): e0247959

76. Sadarangani T, Zhong J, Vora P, Missaelides L. “Advocating Every Single Day” so as not to be forgotten: Factors supporting resiliency in adult day service centers amidst COVID-19-related closures. Journal of Gerontological Social Work. 2021: 1–12.

77. Fazel M, Wheeler J, Danesh J. Prevalence of serious mental disorder in 7000 refugees resettled in western countries: a systematic review. The Lancet. 2005;365(9467): 1309–14.

78. Wu T, Jia X, Shi H, Niu J, Yin X, Xie J, et al. Prevalence of mental health problems during the COVID-19 pandemic: A systematic review and meta-analysis. J Affect Disord. 2021;281: 91–8.

79. Wong SMY, Hui CLM, Wong CSM, Suen YN, Chan SKW, Lee EHM, et al. Prospective Prediction of PTSD and Depressive Symptoms during Social Unrest and COVID-19 using a Brief Online Tool. Psychiatry Research. 2021: 113773.

80. Liu CH, Zhang E, Wong GTF, Hyun S. Factors associated with depression, anxiety, and PTSD symptomatology during the COVID-19 pandemic: Clinical implications for US young adult mental health. Psychiatry research. 2020;290: 113172.

81. Torales J, O’Higgins M, Castaldelli-Maia JM, Ventriglio A. The outbreak of COVID-19 coronavirus and its impact on global mental health. International Journal of Social Psychiatry. 2020;66(4): 317–20.

82. Naik SS, Gowda GS, Shivaprakash P, Subramaniyam BA, Manjunatha N, Muliyala KP, et al. Homeless people with mental illness in India and COVID-19. The Lancet Psychiatry. 2020;7(8): e51–e2.

83. Martin C, Andrés P, Bullón A, Villegas JL, de la Iglesia-Larrad JI, Bote B, et al. COVID pandemic as an opportunity for improving mental health treatments of the homeless people. International Journal of Social Psychiatry. 2020: 20764020950770.

